# The Role of Birth Companionship in Women’s Experiences of Mistreatment during Childbirth and Postpartum Depression

**DOI:** 10.1101/2024.11.21.24317712

**Authors:** Waqas Hameed, Bushra Khan, Bilal Iqbal Avan

## Abstract

**Introduction:** Birth companionship during facility-based childbirth has been recommended by the World Health Organization to improve maternal and newborn outcomes. However, limited evidence exists on how companionship affects women’s experiences of mistreatment during childbirth and their mental health.

**Objectives:** To assess the characteristics of birth companionship during intrapartum care and examine its associations with women’s experiences of mistreatment and postpartum anxiety and depression.

**Methods:** We analysed cross-sectional data on 314 women who gave birth in six public health facilities in rural Sindh, Pakistan. Women were interviewed 42 days postpartum about their childbirth experiences and postpartum anxiety and depression. Multivariable regression models were used to examine associations between birth companionship, mistreatment, and postpartum anxiety/depression. Path analysis explored whether mistreatment mediated the relationship between companionship and postpartum anxiety and depression.

**Results:** Most women (83.1%) had a full-time companion in the labor room, with in-laws (44.6%) being the most common, followed by family members (26.1%) and friends (15.0%). Higher levels of companion support were associated with lower levels of mistreatment, particularly regarding non-confidential care, lack of supportive care, and ineffective communication. Women receiving low companion support were 2.86 times more likely to experience postpartum anxiety and depression compared to those receiving high support. Informational support emerged as the strongest protective factor against both mistreatment and postpartum anxiety/depression. Path analysis revealed that the effect of companion support on postpartum mental health was mediated by experiences of mistreatment during childbirth.

**Conclusion:** Birth companionship, especially informational support, plays a crucial role in reducing mistreatment during childbirth and improving maternal mental health outcomes. These findings underscore the need for supportive policies and health system interventions that actively encourage the engagement of companions from a woman’s personal network during labor and childbirth. Future research should explore strategies to optimize the role of birth companions in promoting respectful maternity care and maternal well-being.

## Introduction

One-third of women perceive their childbirth experience as traumatic[1], a significant event that can profoundly change a woman’s life and leave enduring memories. In low- and middle-income countries (LMICs), about 1 in 5 women suffer from common perinatal mental disorders[2]. These psychological distresses during labour make women uniquely vulnerable to environmental influences such as unfamiliar personnel, medicalized procedures, and other conditions, leading them to prefer the presence of someone familiar and comfortable during labour and childbirth[3]. Yet, health systems in LMICs primarily focus on physical health, often deprioritizing or neglecting social and emotional needs. Unfortunately, women are often faced with disrespectful and abusive behaviours during childbirth in facility-based settings[4]. These woeful experiences are likely to have both short- and long-term adverse effects, such as dissatisfaction with maternity care, feelings of re-traumatization, postpartum depression (PPD), and challenges in maternal-infant bonding[3,5].

The World Health Organization (WHO) has strongly recommended the presence of companions of choice during labour and childbirth in their recent guidelines on maternal and newborn care[6–8]; birth companion is also considered as an integral component of respectful maternity care[9]. A review of qualitative studies highlighted the birth companion could play a diverse tangible supportive roles, that include emotional support (e.g., reassurance and praise), informational support (e.g., updates on labour progress and coping techniques), instrumental support (e.g., massage, encouraging mobility), and advocacy (e.g., helping the woman articulate her wishes to others)[10]. A 2017 Cochrane systematic review showed that supportive care during childbirth either from personal companion or professional team improves a range of maternal and infant outcomes. These include a shorter duration of labour, higher perceived control over birth, lower perceived labour pain, a greater likelihood of spontaneous vaginal birth, and increased maternal satisfaction with childbirth services[3]. Despite promising evidence and recommendations from the WHO, the coverage of companionship during labour and childbirth varies considerably across countries, ranging from 4% to 94%[11].

While several studies have sought to enhance understanding of birth companionship, including general perceptions, associated factors, and its impact on maternal outcomes, limited evidence is available on how companionship affects women’s experiences of mistreatment during childbirth and their mental health. Few studies[12–14] came out recently and all of them consistently showed that presence of birth companion reduces the likely of mistreatment. However, none of them examined the characteristics of birth companionship and its relationship with mistreatment[12–14]. Furthermore, there is research examining the relationship between birth companions and maternal mental health. Most of the existing research has focused on the presence of a personal birth companion rather than the impact of the characteristics of that companionship on mental health[15–18]. This study seeks to address existing gaps by: (a) assessing the characteristics of birth companionship during intrapartum care; (b) examining the associations between birth companionship, women’s experiences of mistreatment, and postpartum anxiety and depression; and (c) exploring whether the relationship between birth companionship and postpartum depression is mediated by women’s experiences of mistreatment.

### Research Question

What are the characteristics of birth companionship during intrapartum care, and how they are associated with women’s experiences of mistreatment and postpartum anxiety and depression?

## Methods

We report on the endline survey data collected between 13^th^ September and 11^th^ December 2021, as part of a larger experimental study conducted in two rural districts of southern Sindh, Pakistan[19]. The main study aimed to promote supportive and respectful maternity care (S-RMC) in six secondary-level public health care facilities. The health facilities provided at least standard basic emergency obstetric and newborn care services. Deliveries ranged from 40 to 300 per month at each facility. The study protocol and the results of primary analysis has been published[19,20]. Briefly, S-RMC intervention targeted skill enhancement of maternity teams and systemic improvements for better governance and accountability in health facilities. It involved comprehensive team training in respectful and psychosocial care, followed by the implementation of practices like vulnerability screening and provision of respectful care and psychosocial support during maternity care. Engagement of birth companion from women’s personal network like mother, sister etc. was an integral component of psychosocial support[21]. The intervention also introduced a feedback system through complaints and exit interviews, and administrative measures like S-RMC data review for performance improvement, staff recognition, and professional development. A mental health first-aider supervised these activities, supported by the research team to ensure effective integration of S-RMC practices[19].

### Data collection and management

A total of 314 women were interviewed. Recruitment was limited to women who had given birth at the health facility (study site) during the period of data collection. These women were initially contacted at health facilities and subsequently interviewed in their homes regarding their childbirth experiences, 42 days after delivery. A consecutive sampling technique was used to recruit participants for the study. All women who delivered at the selected facilities during the data collection period were asked to participate in the survey until the desired sample size was achieved. The distribution of the sample across the health facilities was proportional to their childbirth caseloads. Data collection was carried out simultaneously at all six facilities. Only those women who consented to interviews at both the health facility and their homes six weeks postpartum were included in the survey. Women living outside the study district or in remote rural areas were excluded due to logistical difficulties and potential security concerns for the data collectors. Data collection was conducted electronically using tablets, utilizing Epicollect5 software which included built-in validation checks to maintain data quality. Routine assessments of data quality were performed through simple frequency counts and cross-classification analysis. All participating women provided written informed consent.

### Measures

#### Dependent variables

We used two outcome variables: women’s experiences of mistreatment (continuous scale, 0-100) and postpartum anxiety and depression (binary: no/yes). An open-access, community-based structured instrument, developed by an international group, was adapted to document experiences of mistreatment[22]. We utilized the WHO’s mistreatment framework to develop a composite measure for overall mistreatment and specific types. The overall mistreatment score was derived from 53 binary items, scoring "1" for experienced mistreatment and "0" for none, with total scores ranging from 0 to 53 and linearly transformed to a 0-100 scale. This score reflects the cumulative level of mistreatment during maternity care, where higher scores indicate women experienced greater number of mistreatment manifestations. We applied the same scoring method for each mistreatment type, also scaled from 0 to 100. The symptoms of anxiety and depression was screened using Patient Health Questionnaire – 4 at 42 days postpartum[23]. Standard cut-off was used to create a binary measure.

### Independent variable

Birth companions referred to individuals chosen by a woman from her personal network—such as her husband, mother, or a friend—to provide support during labour and childbirth.

We used two main independent variables: type of birth companion and quality of support provided by the birth companion during the stay in the health facility for childbirth. We created a composite score to reflect quality of companion support based on following nine binary items that were classified into three categories: A) Emotional support: (i) console through touch and reassuring words, (ii) distract by talking about any subject to ease anxiety or pain, (iii) avoid doing or saying anything that may hurt or upset; B) Instrumental support: (i) Help her adopt an alternative position to ease pain, (ii) Maintain privacy, (iii) Assist to ambulate during labour; C) Informational support: (i) encourage and/or remind her of the breathing exercise, (ii) update women about current condition, (iii) support regarding nutritional and medicinal intake.

The overall composite score of quality of companion support was constructed by summing the scores of 9 binary items, where “1” indicates “yes,” and “0” indicates “no”. The total raw score ranged from 0 to 9, where a higher score indicates a higher level of support provided to the women during maternity care. The internal consistency for the overall measure was 0.75. The same procedure was repeated for creating composite scores for each type of support, each ranging from 0 to 3. The overall measure of companionship was categorised into: low (0-5), moderate (6-7), and high (8-9) level of support.

### Statistical analysis

The statistical analyses were performed in multiple steps. First, we used descriptive analysis to describe the characteristics of study population, and level and kind of support provided by the birth companion to women. Second, we used multivariable regression models to determine the relationship between measures of birth companion and mistreatment and postpartum anxiety/depression. In view of the type of outcomes variable (continuous or binary), multivariable linear and logistic regression models were used. All models were adjusted for covariates, including age, order of pregnancy, ethnicity, education, household poverty, level of involvement in household decision-making, mode of birth, and sex of index baby. Finally, we used path analysis to examine whether or not the relationship between birth companionship (continuous) and postpartum anxiety/depression is mediated by experiences of mistreatment during childbirth. We fitted four separate path models: one using the overall measure of companionship and three others for each specific type of companion support. It is important to note that we used discrete variables in these models, with reverse coding to indicate the lack of companion support.

Stata version 18.0 (StataCorp, College Station, TX) was used for all analyses. P-value of <0.05 were considered significant.

### Ethical Approval

The study protocol, the informed consent forms, and other appropriate documents were approved by the Ethics Review Committee of the Aga Khan University (Ref. ID: 2019-1683-5607) and Research Ethics Committee of the London School of Hygiene & Tropical Medicine (Ref. ID: 17886). The study has been registered with clinicaltrials.gov (registration number: NCT05146518).

## Results

Table 1 describes the socio-demographic and reproductive characteristics of the study participants. The mean age of the women was 29.7 (±5.2) and about one-fourth had at least one functional disability. The majority were Sindhi (93.0%) spoken, had no formal education (81.2%), and about half of them were living on less than $1.25 a day. Regarding reproductive characteristics, 84.4% were primigravida, 93.0% received antenatal care, and 86.0% had vaginal births.

**Table 1:**
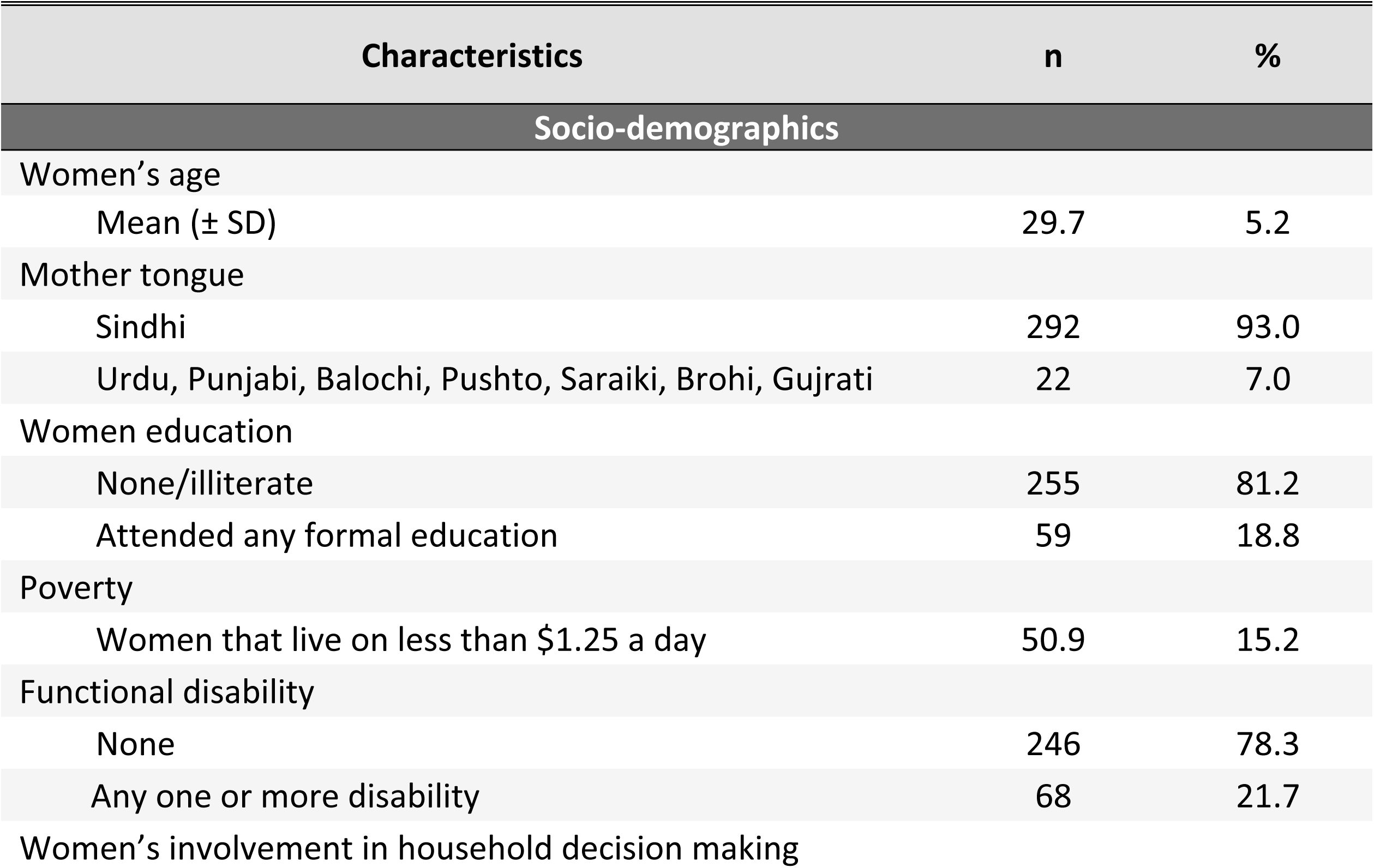
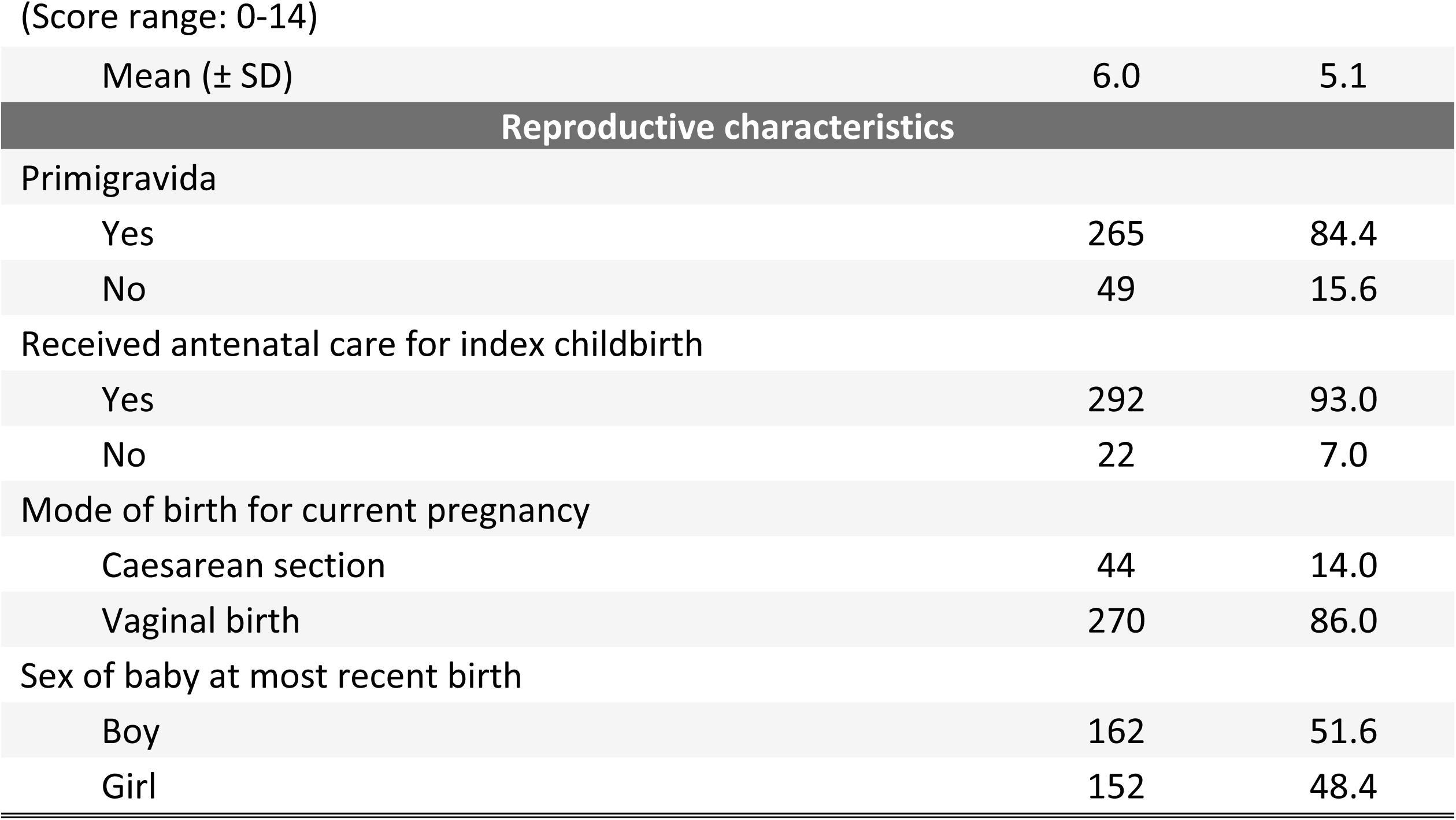
Socio-demographic and reproductive characteristics of study participants.

| Characteristics | n | % |
| --- | --- | --- |
| **Socio-demographics** |  |  |
| Women's age |  |  |
| Mean (± SD) | 29.7 | 5.2 |
| Mother tongue |  |  |
| Sindhi | 292 | 93.0 |
| Urdu, Punjabi, Balochi, Pushto, Saraiki, Brohi, Gujrati | 22 | 7.0 |
| Women education |  |  |
| None/illiterate | 255 | 81.2 |
| Attended any formal education | 59 | 18.8 |
| Poverty |  |  |
| Women that live on less than \$1.25 a day | 50.9 | 15.2 |
| Functional disability |  |  |
| None | 246 | 78.3 |
| Any one or more disability | 68 | 21.7 |
| Women's involvement in household decision making |  |  |
| (Score range: 0-14) |  |  |
| Mean (± SD) | 6.0 | 5.1 |
| **Reproductive characteristics** |  |  |
| Primigravida |  |  |
| Yes | 265 | 84.4 |
| No | 49 | 15.6 |
| Received antenatal care for index childbirth |  |  |
| Yes | 292 | 93.0 |
| No | 22 | 7.0 |
| Mode of birth for current pregnancy |  |  |
| Caesarean section | 44 | 14.0 |
| Vaginal birth | 270 | 86.0 |
| Sex of baby at most recent birth |  |  |
| Boy | 162 | 51.6 |
| Girl | 152 | 48.4 |

Table 2 details the presence and role of personal companions during labour and childbirth. An overwhelming majority (98.7%) had a companion at some point during their hospital stay. Companions were most commonly present after childbirth (98.4%), during labour (93.6%), and during childbirth itself (85.7%). Most women (83.1%) were allowed a companion full-time in the labour room. In the labour room, the majority of women were accompanied by their in-laws (44.6%), followed by family members (26.1%), and friends (15.0%).

**Table 2:**
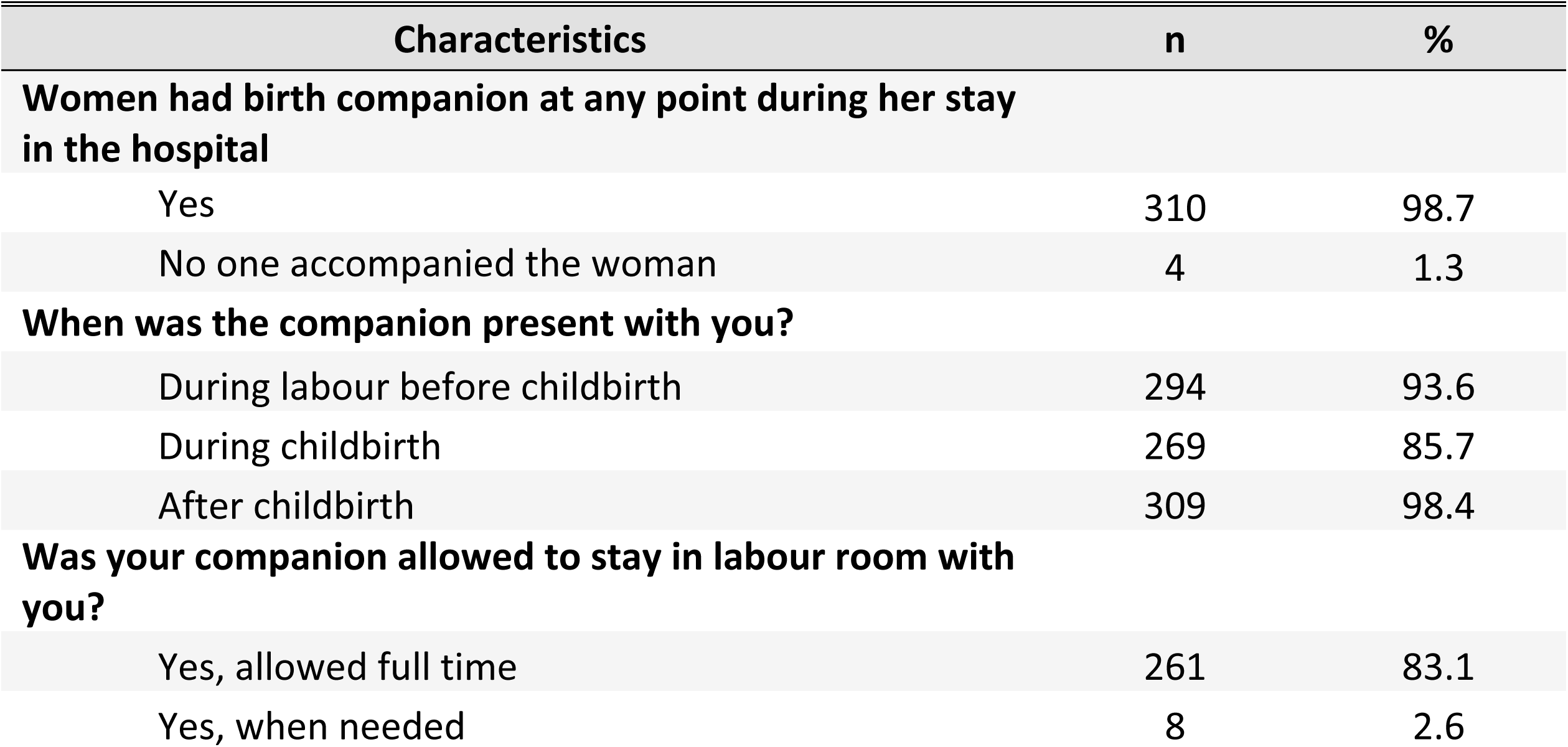
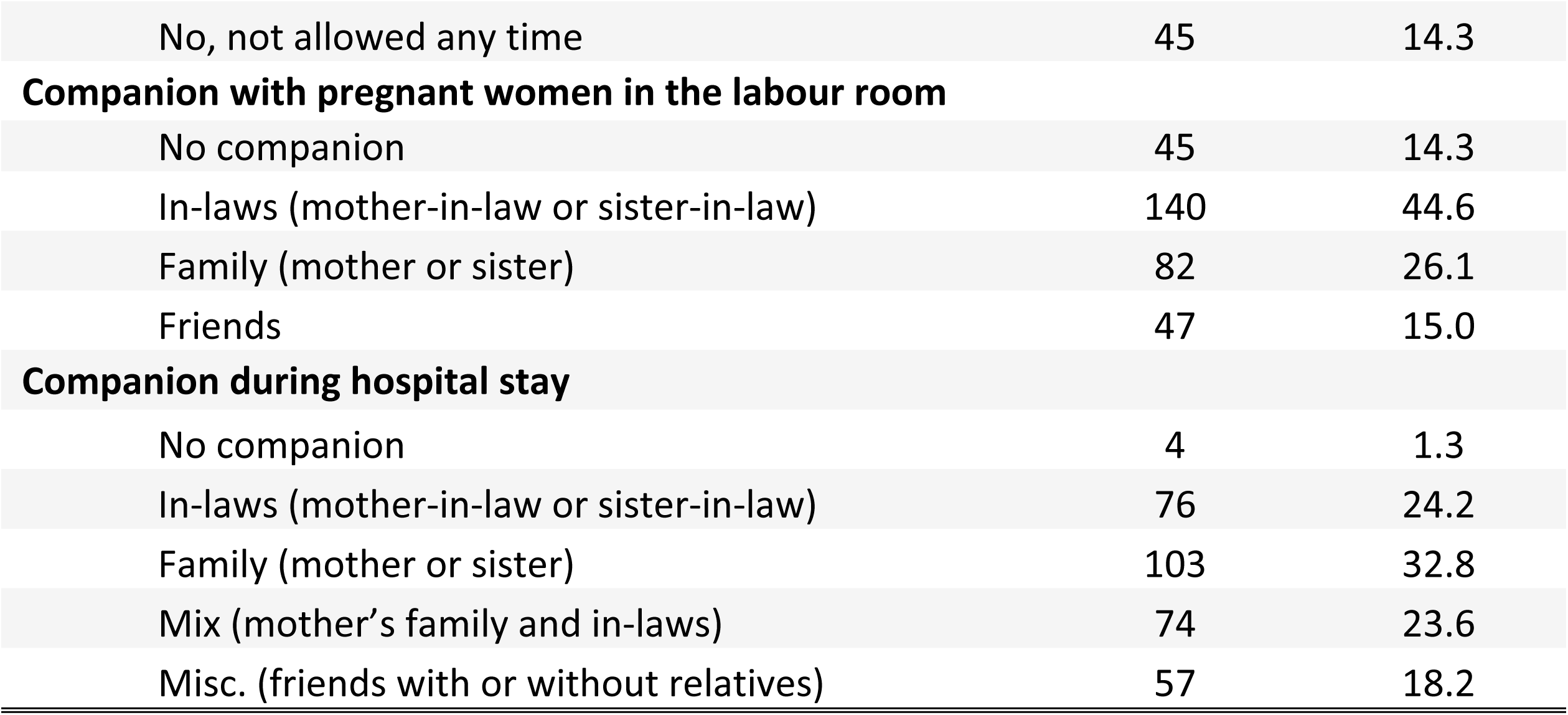
Companion support during labour and childbirth (n=310)

| Characteristics | n | % |
| --- | --- | --- |
| **Women had birth companion at any point during her stay in the hospital** |  |  |
| Yes | 310 | 98.7 |
| No one accompanied the woman | 4 | 1.3 |
| **When was the companion present with you?** |  |  |
| During labour before childbirth | 294 | 93.6 |
| During childbirth | 269 | 85.7 |
| After childbirth | 309 | 98.4 |
| **Was your companion allowed to stay in labour room with you?** |  |  |
| Yes, allowed full time | 261 | 83.1 |
| Yes, when needed | 8 | 2.6 |
| No, not allowed any time | 45 | 14.3 |
| **Companion with pregnant women in the labour room** |  |  |
| No companion | 45 | 14.3 |
| In-laws (mother-in-law or sister-in-law) | 140 | 44.6 |
| Family (mother or sister) | 82 | 26.1 |
| Friends | 47 | 15.0 |
| **Companion during hospital stay** |  |  |
| No companion | 4 | 1.3 |
| In-laws (mother-in-law or sister-in-law) | 76 | 24.2 |
| Family (mother or sister) | 103 | 32.8 |
| Mix (mother's family and in-laws) | 74 | 23.6 |
| Misc. (friends with or without relatives) | 57 | 18.2 |

Figure 1 illustrates that the most common types of support provided by birth companions were: informational (79.6.0%), followed by instrumental (63.4%), and emotional (12.4%). Furthermore, the most commonly provided supports were consoling touch and reassuring words (96.8%), supporting nutritional and medicinal needs (96.8%), and maintained privacy (96.2%). On the contrary, avoid doing or saying anything to the women that may hurt was the lowest with 13.4%. The majority of women received multiple forms of support, with a substantial proportion receiving eight out of nine possible types of support assessed (results not shown).

**Figure 1:**
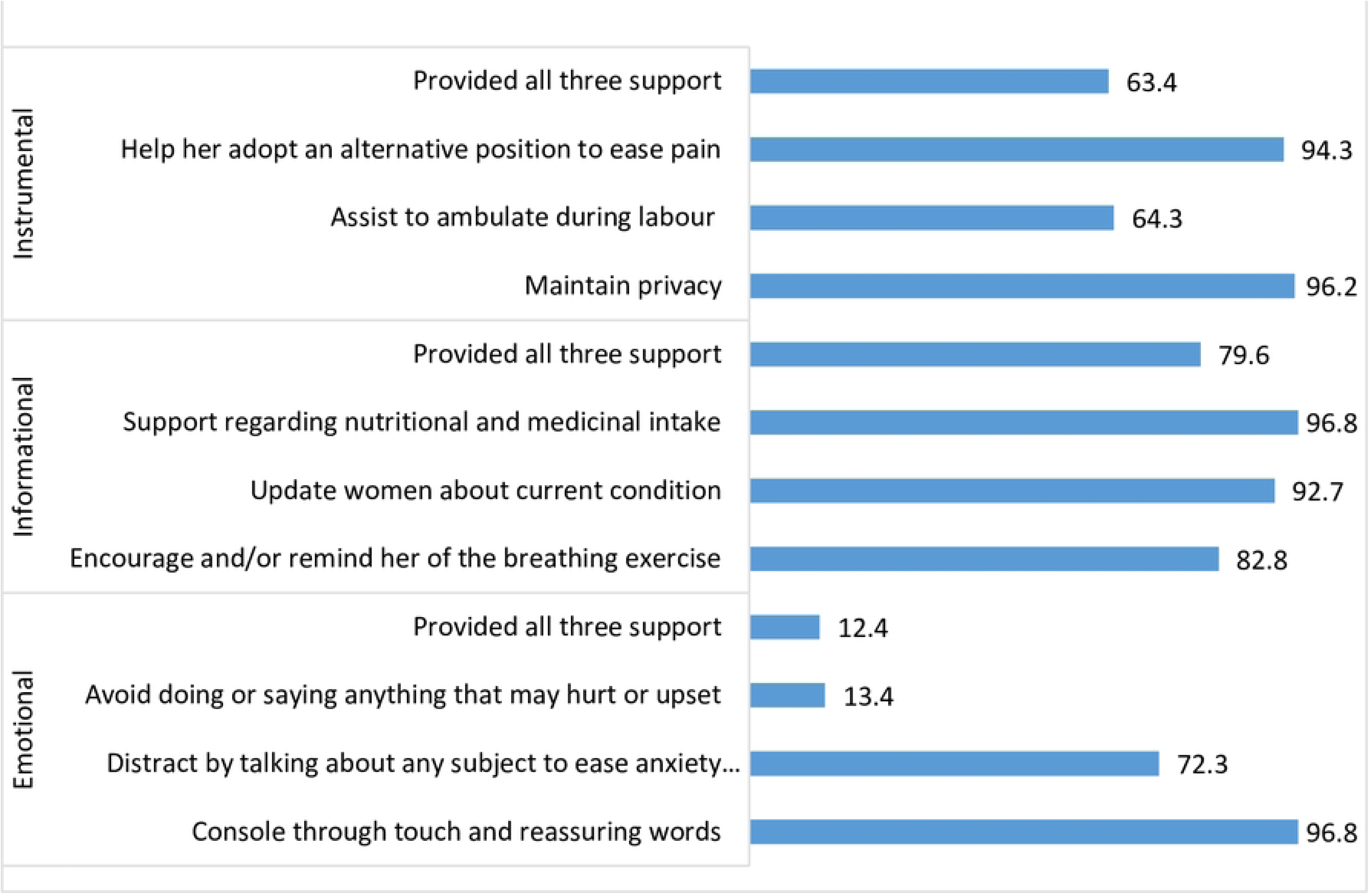
Characteristics of birth companionship.

Table 3 presents the relationship between the level of companion support and women’s experiences of mistreatment during childbirth, as well as postpartum anxiety and depression. Lack of companion support led to a significant increase in women’s experiences of overall mistreatment, lack of professional standards, ineffective communication, non-confidential care, non-inclusive care, and lack of supportive care. Compared with women who received high companion support, the cumulative level of mistreatment was significantly higher among women who received moderate (adjusted β = 3.68, 95% CI [1.25, 6.11]; p < 0.05) and high (adjusted β = 8.19, 95% CI [2.53, 13.85]; p < 0.05) support. Similarly, the level of non-confidential care was significantly higher among women who received moderate companion support (adjusted β = 7.86, 95% CI [3.97, 11.75]; p < 0.01) and even more so among those who received low support (adjusted β = 19.81, 95% CI [11.53, 28.08]; p < 0.01), compared to women with high support. Furthermore, the mean score of ineffective communication was 15.49 units higher (95% CI −0.02, 30.99; p=0.05) among women received low companion support as compared with women receiving high support. The mean score for lack of supportive care substantially increased among women who received moderate companion support (adjusted β = 16.61, 95% CI [11.93, 21.29]; p < 0.01), and this increase was even greater among those with low companion support (adjusted β = 27.32, 95% CI [14.16, 40.49]; p < 0.001), in comparison to women who received high companion support. Women who received low support from their birth companions were 2.86 times (95% CI 1.52 – 5.39) more likely to experience postpartum anxiety and depression compared to those who received high support. The experiences of lack of professional standards was high among women who received low support from their companion as compared with their counterparts. Counterintuitively, we found that a lack of companion support was associated with fewer experiences of non-inclusive care. Women who received low companion support reported lower scores (adjusted β = −10.77, 95% CI [−13.05, −8.49]; p < 0.001) for non-inclusive care compared to those who received high companion support.

**Table 3:** Relationship between companion support and women’s experiences of mistreatment during childbirth and postpartum anxiety and depression.

| Characteristics | Crude<br>$\beta$ (95% CI) | Adjusted<sup>1</sup><br>$\beta$ (95% CI) |
| --- | --- | --- |
| **SUPPORTIVE AND RESPECTFUL MATERNITY CARE (score range: 0-100)** |  |  |
| **Overall mistreatment** |  |  |
| Companion support [Ref: High (Score:8-9)] | 1 | 1 |
| Moderate (Score: 6-7) | 3.57 (0.95 - 6.19)* | 3.68 (1.25 - 6.11)* |
| Low (Score: 0-5) | 8.66 (3.71 - 13.61)** | 8.19 (2.53 - 13.85)* |
| **RESPECTFUL MATERNITY CARE (score range: 0-100)** |  |  |
| **Physical abuse** |  |  |
| Companion support [Ref: High (Score:8-9)] | 1 | 1 |
| Moderate (Score: 6-7) | 1.45 (0.18 - 2.71)* | 1.4 (-0.26 - 3.06) |
| Low (Score: 0-5) | 0.89 (-3.1 - 4.87) | 0.85 (-2.54 - 4.24) |
| **Verbal abuse** |  |  |
| Companion support [Ref: High (Score:8-9)] | 1 | 1 |
| Moderate (Score: 6-7) | 1.35 (-0.71 - 3.41) | 1.42 (-0.36 - 3.2) |
| Low (Score: 0-5) | -0.09 (-3.16 - 2.99) | -0.64 (-6.12 - 4.85) |
| **Lack of professional standards** |  |  |
| Companion support [Ref: High (Score:8-9)] | 1 | 1 |
| Moderate (Score: 6-7) | 0.77 (-3.12 - 4.65) | 1.66 (-1.42 - 4.74) |
| Low (Score: 0-5) | 5.28 (0.03 - 10.54)* | 5.2 (0.47 - 9.93)* |
| **Ineffective communication** |  |  |
| Companion support [Ref: High (Score:8-9)] | 1 | 1 |
| Moderate (Score: 6-7) | 4.19 (-6.56 - 14.94) | 3.93 (-7.05 - 14.9) |
| Low (Score: 0-5) | 16.49 (2.46 - 30.52)* | 15.49 (-0.02 - 30.99)* |
| **Non-confidential care** |  |  |
| Companion support [Ref: High (Score:8-9)] | 1 | 1 |
| Moderate (Score: 6-7) | 8.04 (4.77 - 11.31)** | 7.86 (3.97 - 11.75)** |
| Low (Score: 0-5) | 20.74 (13.62 - 27.85)** | 19.81 (11.53 - 28.08)** |
| **Health system culture & constraints** |  |  |
| Companion support [Ref: High (Score:8-9)] | 1 | 1 |
| Moderate (Score: 6-7) | -1.14 (-11.31 - 9.03) | -0.29 (-6.4 - 5.83) |
| Low (Score: 0-5) | 4.63 (-8.86 - 18.12) | 6.06 (-3.72 - 15.84) |
| **INCLUSIVE CARE (score range: 0-100)** |  |  |
| **Non-inclusive care** |  |  |
| Companion support [Ref: High (Score:8-9)] | 1 | 1 |
| Moderate (Score: 6-7) | -8.37 (-14.64 - [-2.09])* | -8.69 (-14.97 - [-2.41])* |
| Low (Score: 0-5) | -10.01 (-11.95 - [-8.07])*** | -10.77 (-13.05 - [-8.49])*** |
| **Stigma and discrimination** |  |  |
| Companion support [Ref: High (Score:8-9)] | 1 | 1 |
| Moderate (Score: 6-7) | 0.58 (-0.45 - 1.61) | 0.52 (-0.47 - 1.52) |
| Low (Score: 0-5) | -0.17 (-0.45 - 0.1) | -0.41 (-0.83 - 0.01) |
| **SUPPORTIVE CARE (score range: 0-100)** |  |  |
| **Lack of supportive care** |  |  |
| Companion support [Ref: High (Score:8-9)] | 1 | 1 |
| Moderate (Score: 6-7) | 16.94 (12.02 - 21.86)** | 16.61 (11.93 - 21.29)** |
| Low (Score: 0-5) | 30.17 (18.29 - 42.06)*** | 27.32 (14.16 - 40.49)*** |
| **MATERNAL MENTAL HEALTH** |  |  |
| **Postpartum anxiety and depression** |  |  |
|  | **OR (95% CI)** | **aOR (95% CI)** |
| Companion support [Ref: High (Score:8-9)] | 1 | 1 |
| Moderate (Score: 6-7) | 0.94 (0.55 - 1.62) | 0.98 (0.6 - 1.6) |
| Low (Score: 0-5) | 2.81 (1.42 - 5.57)** | 2.86 (1.52 - 5.39)** |

<sup>1</sup>Adjusted for women's age, primigravida, mother tongue, education, household poverty, involvement in household decision making, mode of birth, antenatal care for index birth and sex of index baby
\* p<0.05, \*\* p<0.01, \*\*\* p<0.001

The relationship between the three types of companion support and mistreatment, as well as postpartum anxiety and depression, is detailed in Table 4. Women who received less than three forms of instrumental support reported significantly higher experiences of lack of supportive care (adjusted β = 11.61, 95% CI [3.74, 19.49]; p < 0.05). Similarly, lack of emotional support increased the experiences of lack of supportive care (adjusted β = 11.61, 95% CI [3.74, 19.49]; p < 0.05) and ineffective communication (adjusted β = 8.46, 95% CI [4.81, 12.12]; p < 0.01).

**Table 4:** Relationship between type of companion support and women’s experiences of mistreatment during childbirth and postpartum anxiety and depression.

| Characteristics | Instrumental support |  | Emotional support |  | Informational support |  |
| --- | --- | --- | --- | --- | --- | --- |
| | Crude<br>$\beta$ (95% CI) | Adjusted<sup>1</sup><br>$\beta$ (95% CI) | Crude<br>$\beta$ (95% CI) | Adjusted<sup>1</sup><br>$\beta$ (95% CI) | Crude<br>$\beta$ (95% CI) | Adjusted<sup>1</sup><br>$\beta$ (95% CI) |
| **SUPPORTIVE AND RESPECTFUL MATERNITY CARE (score range: 0-100)** |  |  |  |  |  |  |
| **Overall mistreatment** |  |  |  |  |  |  |
| All three support | 1 | 1 | 1 | 1 | 1 | 1 |
| Less than three support | 2.13 (-0.06 - 4.32) | 1.88 (-0.2 - 3.96) | 3.2 (0.31 - 6.09)* | 2.8 (-0.65 - 6.25) | 8.43 (4.55 - 12.31)** | 8.12 (3.74 - 12.5)** |
| **RESPECTFUL MATERNITY CARE (score range: 0-100)** |  |  |  |  |  |  |
| **Physical abuse** |  |  |  |  |  |  |
| All three support | 1 | 1 | 1 | 1 | 1 | 1 |
| Less than three support | 1 (-0.49 - 2.5) | 0.95 (-0.44 - 2.34) | 0.78 (-1 - 2.56) | 0.96 (-1.05 - 2.96) | 1.34 (-0.46 - 3.14) | 1.27 (-0.28 - 2.81) |
| **Verbal abuse** |  |  |  |  |  |  |
| All three support | 1 | 1 | 1 | 1 | 1 | 1 |
| Less than three support | 0.13 (-1.98 - 2.25) | -0.12 (-1.49 - 1.24) | 0.77 (-4.09 - 5.64) | 0.87 (-5.67 - 7.42) | 2.68 (0.14 - 5.22)* | 2.08 (-1.78 - 5.94) |
| **Lack of professional standards** |  |  |  |  |  |  |
| All three support | 1 | 1 | 1 | 1 | 1 | 1 |
| Less than three support | 1.83 (-1.26 - 4.92) | 1.63 (-1.31 - 4.56) | -2.26 (-10.29 - 5.76) | -1.8 (-8.93 - 5.33) | 5.87 (-0.82 - 12.55) | 6.11 (0.02 - 12.19)* |
| **Ineffective communication** |  |  |  |  |  |  |
| All three support | 1 | 1 | 1 | 1 | 1 | 1 |
| Less than three support | 0.95 (-6.71 - 8.61) | 0.2 (-6.56 - 6.95) | 10.2 (5.22 - 15.17)** | 8.46 (4.81 - 12.12)** | 16.06 (0.69 - 31.43)* | 15.77 (0.06 - 31.48)* |
| **Non-confidential care** |  |  |  |  |  |  |
| All three support | 1 | 1 | 1 | 1 | 1 | 1 |
| Less than three support | 2.08 (-0.99 - 5.16) | 1.44 (-1.82 - 4.69) | 6.85 (0.94 - 12.76)* | 5.43 (-2.58 - 13.45) | 16.83 (12.05 - 21.62)*** | 15.9 (10.02 - 21.78)** |
| **Health system culture & constraints** |  |  |  |  |  |  |
| All three support | 1 | 1 | 1 | 1 | 1 | 1 |
| Less than three support | -0.38 (-8.62 - 7.86) | -0.24 (-7.24 - 6.76) | -2.43 (-12.91 - 8.05) | -0.1 (-3.83 - 3.63) | 3.73 (-5.02 - 12.47) | 6.04 (-2.42 - 14.51) |
| **INCLUSIVE CARE (score range: 0-100)** |  |  |  |  |  |  |
| **Non-inclusive care** |  |  |  |  |  |  |
| All three support | 1 | 1 | 1 | 1 | 1 | 1 |
| Less than three support | -4.17 (-9.24 - 0.9) | -4.08 (-8.81 - 0.64) | -4.55 (-12.48 - 3.39) | -4.86 (-13.95 - 4.24) | -7.29 (-10.64 - -3.94)** | -7.77 (-11.16 - -4.37)** |
| **Stigma and discrimination** |  |  |  |  |  |  |
| All three support | 1 | 1 | 1 | 1 | 1 | 1 |
| Less than three support | 0.41 (-0.85 - 1.66) | 0.37 (-0.78 - 1.52) | 0 (-0.55 - 0.55) | -0.1 (-0.91 - 0.72) | 0.66 (-1.34 - 2.67) | 0.38 (-1.16 - 1.92) |
| **SUPPORTIVE CARE (score range: 0-100)** |  |  |  |  |  |  |
| **Lack of supportive care** |  |  |  |  |  |  |
| All three support | 1 | 1 | 1 | 1 | 1 | 1 |
| Less than three support | 12.91 (4.67 - 21.14)* | 11.61 (3.74 - 19.49)* | 14.91 (9.77 - 20.05)** | 11.46 (3.49 - 19.42)* | 28.69 (21.37 - 36)*** | 25.49 (18 - 32.97)*** |
| **MATERNAL MENTAL HEALTH** |  |  |  |  |  |  |
| **Postpartum anxiety and depression** |  |  |  |  |  |  |
| All three support | 1 | 1 | 1 | 1 | 1 | 1 |
| Less than three support | 1.55 (0.89 - 2.71) | 1.64 (0.97 - 2.8) | 0.76 (0.35 - 1.65) | 0.74 (0.32 - 1.67) | 2.19 (1.19 - 4.04)* | 2.12 (1.03 - 4.35)* |

<sup>1</sup>Adjusted for women's age, primigravida, mother tongue, education, household poverty, involvement in household decision making, mode of birth, antenatal care for index birth and sex of index baby
\* p<0.05, \*\* p<0.01, \*\*\* p<0.001

Lack of informational support was associated with overall mistreatment (adjusted β = 8.12, 95% CI [3.74, 12.5]; p < 0.01), including specific types of mistreatment: lack of professional standards (adjusted β = 6.11, 95% CI [0.02, 12.19]; p < 0.05), ineffective communication (adjusted β = 15.77, 95% CI [0.06, 31.48]; p < 0.05), non-confidential care (adjusted β = 15.9, 95% CI [10.02, 21.78]; p < 0.01), and lack of supportive care (adjusted β = 25.49, 95% CI [18.0, 32.97]; p < 0.001). Notably, the odds of postpartum anxiety and depression were 2.21 times higher (95% CI [1.03, 4.35]) among women who did not receive all three forms of informational support from their companion compared to those who did.

Results from the mediation analysis indicate that the effect of companion support on postpartum anxiety and depression is mediated by experiences of mistreatment during childbirth, with every unit reduction in companion support the average cumulative level of overall mistreatment increased by 1.367 units (95% CI 0.11-2.62; p = 0.038). Furthermore, a unit increase in the cumulative level of overall mistreatment is associated with a 4% increase in the likelihood of postpartum anxiety and depression (OR = 1.041, 95% CI 1.01, 1.08; p = 0.034). However, the direct effect of companion support on postpartum depression anxiety and depression was only marginally significant (OR=1.51, p=0.061) (figure 2A). Regarding the type of companion support, emotional support’s effect on postpartum anxiety and depression was mediated through experiences of mistreatment, while the direct relationship was not significant (OR = 1.083, p = 0.810) (Figure 2B). In contrast, instrumental support had a direct, significant association with postpartum anxiety and depression, but its indirect effects were not significant (Figure 2C). Notably, informational support had a significant impact on postpartum anxiety and depression through both direct and indirect pathways (Figure 2D).

**Figure 2A-D:**
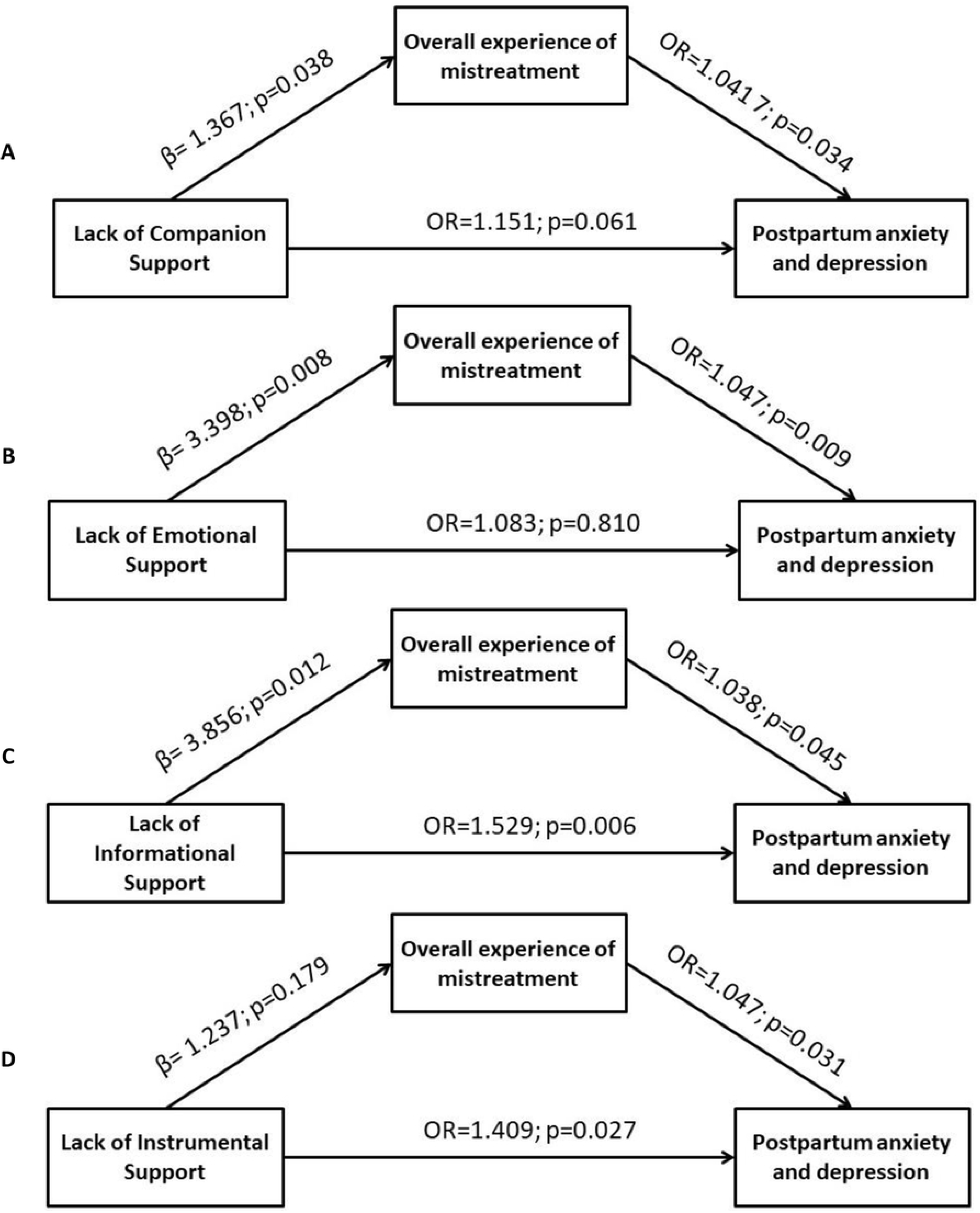
Links between companionship, mistreatment, and postpartum anxiety and depression.

## Discussion

Despite the recommendation from the WHO of allowing women to choose a personal birth companion, the policy is not widely implemented across many LMICs[8]. While research shows that having a personal birth companion can reduce mistreatment during childbirth, there is limited evidence on how the characteristics of the companionship affect mistreatment and maternal mental health. We examined the characteristics of support provided by birth companions and their association with women’s experiences of mistreatment and postpartum anxiety and depression.

Our study found that the vast majority of women were accompanied by a personal companion during labour, childbirth, and post-delivery, typically in-laws and family members who provided substantial informational, emotional, and practical support. Higher levels of companion support were significantly associated with reduced mistreatment, particularly in communication, respect for confidentiality, and supportive care. Women who received moderate to high levels of support experienced less mistreatment and overall better care compared to those who received low support. Additionally, greater companion support was linked to reduced postpartum anxiety and depression, primarily due to a decrease in mistreatment experiences.

The presence of birth companions in our study is higher compared to other South Asian countries[24], including Bangladesh (55.5%)[25] and India (73%)[26]. However, it aligns with findings from a previous study conducted in Pakistan[27]. A likely reason for this higher presence is that our analysis is based on a post-intervention survey where birth companionship was a core component of the intervention[19,21,28]. Moreover, companion engagement was perceived as an effective task-sharing strategy that allowed maternity service providers to offer continuous support, thereby reducing their workload[28–30]. In our study, the most common companions were in-laws and other family members, excluding husbands. Although husbands were present at the hospital, they were not permitted in maternity wards, including labour rooms, due to restrictive policies[31], socio-cultural norms[32,33], and privacy concerns for other birthing women, as public hospitals often manage multiple women in large labour rooms[33].

Consistent with other studies[12,14,34,35], we observed a strong relationship between birth companionship and reduced mistreatment. Specifically, experiences of non-confidential care, ineffective communication, and lack of supportive care were significantly reduced. Of the three different types of companion support, informational support emerged as a stronger determinant of the experiences of mistreatment and postpartum anxiety and depression. Qualitative studies also reveal that companions play a crucial role in providing physical, informational, and emotional support to birthing women[10]. For instance, companions help bridge communication gaps by explaining technical jargon used by healthcare providers and providing timely information about labour progress. They also assist in maintaining privacy during physical examinations[14,32] and are well-positioned to offer psychosocial support, understanding the emotional needs of birthing women[28,36]. In terms of informational support, two psychological factors—women’s internal locus of control and increased trust in their companion—may explain the link between informational support, mistreatment, and postpartum depression. Labour and childbirth can be distressing and create uncertainty. Informational support, which provides facts, knowledge, or advice, can alleviate this[37]. When companions offer updates on the mother’s health, it may boost her confidence and internal locus of control[38,39], fostering a sense of control and encouraging active participation in the birthing process. Additionally, informational support may strengthen women’s trust in their companions, reassuring them that they have someone connected with the healthcare staff[32]. This support could help prevent the risk of mistreatment and hence postnatal anxiety and depression.

Our study found a statistically significant relationship between birth companionship and reduced postpartum anxiety and depression, mediated by experiences of mistreatment. Notably, the path analysis further signifies the importance of informational support from companions which has a both direct and indirectly effect postpartum anxiety and depression. These findings emphasize the critical role of birth companions not only in addressing mistreatment but also in enhancing maternal mental health. The association between birth companionship and mistreatment aligns with findings from other studies[40–42]. While limited evidence exists on the exact pathways through which companionship improves mental health, a plausible explanation is that support during facility-based childbirth acts as a buffer against psychological stressors[43], reducing stress hormones and improving labour physiology and emotional well-being[44–46]. This support positively influences the fetopelvic relationship, facilitating the birthing process[47], decreasing mistreatment, reducing medical interventions, and lowering the risk of complications. Consequently, companion support leads to better birthing outcomes, greater maternal satisfaction, and improved postpartum mental health. Findings from our pathway analysis further substantiate the proposed mechanism linking companionship with maternal mental health.

Our study has several strengths and limitations. Among the strengths, we adapted validated tools to measure mistreatment and used a locally validated tool to assess postpartum anxiety and depression. In addition, the inclusion of pathway analysis adds unique value by explaining the mechanisms linking companionship with maternal mental health. The measure of companionship was comprehensive, based on nine items capturing various types of support identified in a recently published systematic review. The study’s limitations include its cross-sectional design, which restricts us from drawing causal inferences about the relationship between birth companionship and maternal mental health. Conducted in secondary-level hospitals in southern Sindh, the findings may have limited generalizability to other settings. Although we developed a comprehensive measure of birth companionship, it has not been validated. The analysis is based on data from the endline survey of a larger experimental study in which birth companionship was a key component of the intervention. Therefore, estimates of the increased presence and support of companions should be interpreted cautiously when compared to observational studies.

## Conclusion

Our study findings indicate a strong positive relationship between birth companionship and women’s experiences of mistreatment, particularly regarding non-confidential care, lack of supportive care, and ineffective communication. We also found that the impact of companionship support on postpartum depression is mediated by mistreatment. Of the three types of companion support, informational support emerged as a stronger determinant that has a protective effect on both experiences of mistreatment and postpartum anxiety and depression. These findings highlight the critical role of personal companions during facility-based childbirth in reducing mistreatment and improving maternal mental health. There is a need for supportive policies and health system interventions that actively encourage the engagement of companions from a woman’s personal network during labour and childbirth.

## Data Availability

Data will be available upon request

## References

1. Soet JE, Brack GA, DiIorio C. Prevalence and predictors of women’s experience of psychological trauma during childbirth. Birth. 2003;30: 36–46. doi:10.1046/j.1523-536x.2003.00215.x

2. Woody CA, Ferrari AJ, Siskind DJ, Whiteford HA, Harris MG. A systematic review and meta-regression of the prevalence and incidence of perinatal depression. Journal of Affective Disorders. 2017;219: 86–92. doi:10.1016/j.jad.2017.05.003

3. Bohren MA, Hofmeyr GJ, Sakala C, Fukuzawa RK, Cuthbert A. Continuous support for women during childbirth. Cochrane Database Syst Rev. 2017;2017: CD003766. doi:10.1002/14651858.CD003766.pub6

4. Bohren MA, Vogel JP, Hunter EC, Lutsiv O, Makh SK, Souza JP, et al. The Mistreatment of Women during Childbirth in Health Facilities Globally: A Mixed-Methods Systematic Review. PLOS Medicine. 2015;12: e1001847. doi:10.1371/journal.pmed.1001847

5. Bohren MA, Hunter EC, Munthe-Kaas HM, Souza JP, Vogel JP, Gülmezoglu AM. Facilitators and barriers to facility-based delivery in low- and middle-income countries: a qualitative evidence synthesis. Reproductive Health. 2014;11: 71. doi:10.1186/1742-4755-11-71

6. World Health Organization. WHO recommendations: intrapartum care for a positive childbirth experience. [cited 11 Apr 2023]. Available: https://www.who.int/publications-detail-redirect/9789241550215

7. World Health Organization. WHO guide for integration of perinatal mental health in maternal and child health services. Geneva: World Health Organization; 2022. Available: https://www.who.int/publications-detail-redirect/9789240057142

8. World Health Organization. Companion of choice during labour and childbirth for improved quality of care. World Health Organization; 2020. Available: https://www.who.int/publications/i/item/WHO-SRH-20.13

9. Shakibazadeh E, Namadian M, Bohren M, Vogel J, Rashidian A, Nogueira Pileggi V, et al. Respectful care during childbirth in health facilities globally: a qualitative evidence synthesis. BJOG: An International Journal of Obstetrics & Gynaecology. 2018;125: 932–942. doi:10.1111/1471-0528.15015

10. Bohren MA, Berger BO, Munthe-Kaas H, Tunçalp Ö. Perceptions and experiences of labour companionship: a qualitative evidence synthesis. Cochrane Database Syst Rev. 2019;3: CD012449. doi:10.1002/14651858.CD012449.pub2

11. Bohren MA, Hazfiarini A, Vazquez Corona M, Colomar M, De Mucio B, Tunçalp Ö, et al. From global recommendations to (in)action: A scoping review of the coverage of companion of choice for women during labour and birth. PLOS Glob Public Health. 2023;3: e0001476. doi:10.1371/journal.pgph.0001476

12. Diamond-Smith N, Sudhinaraset M, Melo J, Murthy N. The relationship between women’s experiences of mistreatment at facilities during childbirth, types of support received and person providing the support in Lucknow, India. Midwifery. 2016;40: 114–123. doi:10.1016/j.midw.2016.06.014

13. Maung TM, Mon NO, Mehrtash H, Bonsaffoh KA, Vogel JP, Aderoba AK, et al. Women’s experiences of mistreatment during childbirth and their satisfaction with care: findings from a multicountry community-based study in four countries. BMJ Global Health. 2022;5: e003688. doi:10.1136/bmjgh-2020-003688

14. Wahdan Y, Abu-Rmeileh NME. The association between labor companionship and obstetric violence during childbirth in health facilities in five facilities in the occupied Palestinian territory. BMC Pregnancy and Childbirth. 2023;23: 566. doi:10.1186/s12884-023-05811-2

15. Ghanbari-Homayi S, Fardiazar Z, Meedya S, Mohammad-Alizadeh-Charandabi S, Asghari-Jafarabadi M, Mohammadi E, et al. Predictors of traumatic birth experience among a group of Iranian primipara women: a cross sectional study. BMC Pregnancy Childbirth. 2019;19: 182. doi:10.1186/s12884-019-2333-4

16. Theme Filha MM, Ayers S, da Gama SGN, Leal M do C. Factors associated with postpartum depressive symptomatology in Brazil: The Birth in Brazil National Research Study, 2011/2012. J Affect Disord. 2016;194: 159–167. doi:10.1016/j.jad.2016.01.020

17. Ip WY. Chinese husbands’ presence during labour: a preliminary study in Hong Kong. Int J Nurs Pract. 2000;6: 89–96. doi:10.1046/j.1440-172x.2000.00187.x

18. Gankanda WI, Gunathilake IAGMP, Kahawala NL, Ranaweera AKP. Prevalence and associated factors of post-traumatic stress disorder (PTSD) among a cohort of Srilankan post-partum mothers: a cross-sectional study. BMC Pregnancy and Childbirth. 2021;21: 626. doi:10.1186/s12884-021-04058-z

19. Avan BI, Hameed W, Khan B, Asim M, Saleem S, Siddiqi S. Promoting Supportive and Respectful Maternity Care in Public Health Facilities in Sindh, Pakistan: A Theory-Informed Health System Intervention. Glob Health Sci Pract. 2023;11: e2200513. doi:10.9745/GHSP-D-22-00513

20. Avan BI, Hameed W, Khan B, Asim M, Saleem S, Siddiqi S. Inclusive, supportive and dignified maternity care (SDMC)—Development and feasibility assessment of an intervention package for public health systems: A study protocol. PLOS ONE. 2022;17: e0263635. doi:10.1371/journal.pone.0263635

21. Khan B, Hameed W, Avan BI. Psychosocial support during childbirth: Development and adaptation of WHO’s Mental Health Gap Action Programme (mhGAP) for maternity care settings. PLoS One. 2023;18: e0285209. doi:10.1371/journal.pone.0285209

22. Bohren MA, Vogel JP, Fawole B, Maya ET, Maung TM, Baldé MD, et al. Methodological development of tools to measure how women are treated during facility-based childbirth in four countries: labor observation and community survey. BMC Med Res Methodol. 2018;18: 132. doi:10.1186/s12874-018-0603-x

23. Kroenke K, Spitzer RL, Williams JBW, Löwe B. An ultra-brief screening scale for anxiety and depression: the PHQ-4. Psychosomatics. 2009;50: 613–621. doi:10.1176/appi.psy.50.6.613

24. World Health Organization. Sexual, reproductive, maternal, newborn, child and adolescent health: policy survey, 2018-2019: summary report. Geneva, Switzerland: World Health Organization; 2020. Available: https://iris.who.int/bitstream/handle/10665/331847/9789240004092-eng.pdf?sequence=1

25. Perkins J, Rahman AE, Mhajabin S, Siddique AB, Mazumder T, Haider MR, et al. Humanised childbirth: the status of emotional support of women in rural Bangladesh. Sex Reprod Health Matters. 27: 228–247. doi:10.1080/26410397.2019.1610277

26. Afulani PA, Phillips B, Aborigo RA, Moyer CA. Person-centred maternity care in low-income and middle-income countries: analysis of data from Kenya, Ghana, and India. Lancet Glob Health. 2019;7: e96–e109. doi:10.1016/S2214-109X(18)30403-0

27. Agha S, Fitzgerald L, Fareed A, Rajbhandari P, Rahim S, Shahid F, et al. Quality of labor and birth care in Sindh Province, Pakistan: Findings from direct observations at health facilities. PLoS One. 2019;14: e0223701. doi:10.1371/journal.pone.0223701

28. Avan BI, Hameed W, Khan B, Asim M, Saleem S, Siddiqi S. Understanding the Mechanisms of Change in the Supportive and Respectful Maternity Care Intervention in Sindh, Pakistan: Provider Perspectives. Global Health: Science and Practice. 2023 [cited 18 Dec 2023]. doi:10.9745/GHSP-D-23-00216

29. Sarwal T, Sarwal Y, Tyagi S, Sarwal R. Opinion of Health Care Providers on Birth Companions in Obstetrics Department of a Tertiary Care Hospital in North India. medRxiv; 2021. p. 2021.06.24.21259462. doi:10.1101/2021.06.24.21259462

30. Kabakian-Khasholian T, Bashour H, El-Nemer A, Kharouf M, Elsheikh O, Labour Companionship Study Group. Implementation of a labour companionship model in three public hospitals in Arab middle-income countries. Acta Paediatr. 2018;107 Suppl 471: 35–43. doi:10.1111/apa.14540

31. Maknojia A, Malik A. Why can’t I have a choice of companion during labor? Barriers to implementation of companion presence. Journal of Asian Midwives (JAM). 2021;8: 39–45.

32. Asim M, Hameed W, Khan B, Saleem S, Avan BI. Applying the COM-B Model to Understand the Drivers of Mistreatment During Childbirth: A Qualitative Enquiry Among Maternity Care Staff. Global Health: Science and Practice. 2023;11. doi:10.9745/GHSP-D-22-00267

33. Hameed W, Khan B, Siddiqi S, Asim M, Avan BI. Health system bottlenecks hindering provision of supportive and dignified maternity care in public health facilities. PLOS Global Public Health. 2022;2: e0000550. doi:10.1371/journal.pgph.0000550

34. Balde MD, Nasiri K, Mehrtash H, Soumah A-M, Bohren MA, Diallo BA, et al. Labour companionship and women’s experiences of mistreatment during childbirth: results from a multi-country community-based survey. BMJ Global Health. 2022;5: e003564. doi:10.1136/bmjgh-2020-003564

35. Hajizadeh K, Vaezi M, Meedya S, Mohammad Alizadeh Charandabi S, Mirghafourvand M. Prevalence and predictors of perceived disrespectful maternity care in postpartum Iranian women: a cross-sectional study. BMC Pregnancy Childbirth. 2020;20: 463. doi:10.1186/s12884-020-03124-2

36. Downe S, Finlayson K, Oladapo O, Bonet M, Gülmezoglu AM. What matters to women during childbirth: A systematic qualitative review. PLOS ONE. 2018;13: e0194906. doi:10.1371/journal.pone.0194906

37. Ko H-C, Wang L-L, Xu Y-T. Understanding the Different Types of Social Support Offered by Audience to A-List Diary-Like and Informative Bloggers. Cyberpsychology, Behavior and Social Networking. 2013;16: 194. doi:10.1089/cyber.2012.0297

38. Sandler IN, Lakey B. Locus of control as a stress moderator: The role of control perceptions and social support. Am J Commun Psychol. 1982;10: 65–80. doi:10.1007/BF00903305

39. Moshki M, Cheravi K. Relationships among depression during pregnancy, social support and health locus of control among Iranian pregnant women. Int J Soc Psychiatry. 2016;62: 148–155. doi:10.1177/0020764015612119

40. Salehi A, Fahami F, Beigi M. The effect of presence of trained husbands beside their wives during childbirth on women’s anxiety. Iran J Nurs Midwifery Res. 2016;21: 611–615. doi:10.4103/1735-9066.197672

41. Shahbazi Sighaldeh S, Azadpour A, Vakilian K, Rahimi Foroushani A, Vasegh Rahimparvar SF, Hantoushzadeh S. Comparison of maternal outcomes in caring by Doula, trained lay companion and routine midwifery care. BMC Pregnancy and Childbirth. 2023;23: 765. doi:10.1186/s12884-023-05987-7

42. Sapkota S, Kobayashi T, Takase M. Impact on perceived postnatal support, maternal anxiety and symptoms of depression in new mothers in Nepal when their husbands provide continuous support during labour. Midwifery. 2013;29: 1264–1271. doi:10.1016/j.midw.2012.11.010

43. Hodnett ED, Lowe NK, Hannah ME, Willan AR, Stevens B, Weston JA, et al. Effectiveness of nurses as providers of birth labor support in North American hospitals: a randomized controlled trial. JAMA. 2002;288: 1373–1381. doi:10.1001/jama.288.11.1373

44. Buckley SJ. Executive Summary of Hormonal Physiology of Childbearing: Evidence and Implications for Women, Babies, and Maternity Care. J Perinat Educ. 2015;24: 145–153. doi:10.1891/1058-1243.24.3.145

45. Lederman RP, Lederman E, Work BA, McCann DS. The relationship of maternal anxiety, plasma catecholamines, and plasma cortisol to progress in labor. Am J Obstet Gynecol. 1978;132: 495–500. doi:10.1016/0002-9378(78)90742-1

46. Lederman E, Lederman RP, Work BA, McCann DS. Maternal psychological and physiologic correlates of fetal-newborn health status. American Journal of Obstetrics and Gynecology. 1981;139: 956–958. doi:10.1016/0002-9378(81)90967-4

47. Hofmeyr GJ, Nikodem VC, Wolman WL, Chalmers BE, Kramer T. Companionship to modify the clinical birth environment: effects on progress and perceptions of labour, and breastfeeding. Br J Obstet Gynaecol. 1991;98: 756–764. doi:10.1111/j.1471-0528.1991.tb13479.x

